# State-Transition Dynamics in the HR–Deceleration Capacity Feature Space: Passive Assessment of Autonomic Dysfunction from 24-Hour Heart Rate

**DOI:** 10.64898/2026.09.18.26363446

**Authors:** Lixing Deng, Xingwu Tong, Kuixu Wang, Xingran Cui

**Affiliations:** Center for Nonlinear Dynamics in Medicine School of Biological Science and Medical Engineering Southeast University, Nanjing, China

**Keywords:** Autonomic Nervous System, State-Transition Dynamics, Deceleration Capacity, Long-Term HRV

## Abstract

Autonomic dysfunction is a critical predictor of mortality in aging, Type 2 Diabetes (T2DM), and Congestive Heart Failure (CHF). Active clinical tests such as the Valsalva maneuver and tilt table impose procedural burdens, while standard HRV metrics cannot distinguish transient physiological states from underlying regulatory function. We propose *HR-DC State-Transition Dynamics*, a passive framework that maps 24-hour RR intervals onto a Heart Rate vs. Deceleration Capacity (HR-DC) feature space and partitions it into four physiological states using a Gaussian Mixture Model. An *Onset-Track-Stabilize* protocol then extracts directed state-transition trajectories, yielding three biomarkers (transition efficiency *η*, directional complexity *H*, and tortuosity *τ*) for both Relaxation and Excitation streams. In a longitudinal pilot (*N* = 10, recordings on Monday/Wednesday/Saturday), the three metrics showed high test-retest reliability (ICC *>* 0.85) across days with markedly different activity levels. In a six-group cross-sectional cohort (*N* = 209), an SVM distinguished healthy elderly from pathological subjects with AUC = 0.85, and the metrics correlated with age (*R*^2^ = 0.33). In a clinical subset (*N* = 26) with concurrent active reflex testing, Relaxation metrics predicted Valsalva Ratio (*R*^2^ = 0.58) and Excitation metrics predicted Tilt SBP drop (*R*^2^ = 0.62), indicating that the framework can serve as a passive surrogate for standard autonomic reflex tests.

## I. Introduction

The integrity of the Autonomic Nervous System (ANS) is paramount for cardiovascular homeostasis. Autonomic dysfunction is a known precursor to mortality in Type 2 Diabetes (T2DM) and Congestive Heart Failure (CHF) [1]. The clinical gold standards for ANS assessment, the Valsalva Maneuver and Head-Up Tilt Test, require active patient cooperation [2] and are therefore difficult to apply in geriatric or frail populations. Clinical attention has consequently shifted toward Heart Rate Variability (HRV). Short-term recordings are susceptible to transient confounders [3], and 24-hour Holter-based metrics such as SDNN and LF/HF ratio conflate two distinct phenomena: transient state changes and underlying regulatory function. Low variability may reflect either normal rest or loss of regulatory range due to pathology [4], making it impossible from static metrics alone to distinguish a healthy resting system from one trapped in a low-variability attractor [5].

Several approaches have attempted to recover temporal or geometric information from long-term recordings. Poincaré plot analysis captures beat-to-beat variability geometry within a single-variable delay embedding [5], and DC has been computed over shorter windows to track circadian variation [4], but both remain univariate and cannot jointly represent metabolic load and vagal reserve. No prior work has tracked HR and time-resolved DC as a two-dimensional trajectory or extracted directed state-transition metrics from that joint space.

We hypothesize that healthy autonomic regulation requires the ability to shift between cardiovascular operating points in response to physiological demand, and propose *HR-DC State-Transition Dynamics* to quantify this capacity. The ANS is modeled as a system oscillating between Relaxation (recovery) and Excitation (mobilization) regimes, and directed state transitions are separated from random fluctuations. The contributions of this work are as follows:

- A noise-robust feature space model that employs adaptive, subject-specific partitioning to segment the HR-DC space into four physiological states (*S*_1_–*S*_4_) and a Noise Area.
- A bidirectional Onset-Track-Stabilize protocol extracting six biomarkers that quantify the efficiency and quality of functional state transitions.
- Validation via a longitudinal robustness study, a large multi-cohort classification experiment, and comparison against active clinical reflex benchmarks.

## II. Methods

### A. Study Population and Data Acquisition

This study aggregated data from multiple sources, including open-access databases on PhysioNet [6], previously published datasets collected by our center (SEU), and a newly collected longitudinal validation set. The final cohort (*N* = 209) was stratified into six groups (Table I). Sources differ in sampling rate (128 Hz–1 kHz or RR-only); all ECG signals were resampled to 250 Hz. Healthy controls (Groups A–C) are younger than pathological groups (D–F), reflecting typical open-access database composition. All pathological comparisons were therefore made exclusively against the age-matched Healthy Elderly cohort (Group C); all pathological groups showed no significant age difference from Group C (*p >* 0.05).

**TABLE 1.** Demographics and Data Source Composition.

| Grp | Description | N | Age (yr) | Data Source & Original Hz |
| --- | --- | --- | --- | --- |
| A | Healthy Young | 35 | 24 ± 4 | SEU-HS [7] (20, 500Hz), nsrdb (8, 128Hz), nsr2db (2, RR), rr-healthy (5, RR) |
| B | Healthy Mid | 22 | 42 ± 6 | nsrdb (10, 128Hz), nsr2db (6, RR), rr-healthy (6, RR) |
| C | Healthy Elderly | 47 | 68 ± 7 | nsr2db (46, RR), rr-healthy (1, RR) |
| D | HTN | 39 | 71 ± 8<sup>ns</sup> | shareedb (128Hz)* |
| E | T2DM | 36 | 69 ± 9<sup>ns</sup> | SEU-T2DM [12] (22, 512Hz), cded (14, 1kHz) |
| F | CHF | 30 | 72 ± 10<sup>ns</sup> | chfdb (19, 250Hz), chf2db (11, RR) |
| V1 | Longitudinal Set | 10 | 24 ± 2 | Self-collected (250Hz, 3 days) |
| V2 | Clinical Set | 26 | 65 ± 11 | cded (500Hz, ABP+ECG) |

Data Format: Source (Sample Size $n$ , Sampling Rate).
\*Randomly undersampled. <sup>ns</sup> No sig. age diff vs Grp C ( $p > 0.05$ ).
RR: Beat-interval only datasets (raw ECG are not available).

- Healthy Controls (Groups A–C): Data were sourced from the SEU Healthy Student (SEU-HS) dataset [7], MIT-BIH Normal Sinus Rhythm Database (`nsrdb`) [8], Normal Sinus Rhythm RR Interval Database (`nsr2db`) [9], and the RR Interval Time Series from Healthy Subjects (`rr-interval-healthy`) [10].
- Pathology Groups (Groups D–F):
  - Hypertension (HTN): Randomly undersampled (*n* = 39) from the Smart Health for Assessing the Risk of Events via ECG Database (`shareedb`) [11] to mitigate class imbalance.
  - Type 2 Diabetes (T2DM): Combined the SEU T2DM dataset (SEU-T2DM) [12] with the CAST RR Interval Sub-study Database (`cded`) [13].
  - Congestive Heart Failure (CHF): Combined the BIDMC Congestive Heart Failure Database (`chfdb`) [14] and the Congestive Heart Failure RR Interval Database (`chf2db`) [15].
- Validation Sets: (1) A newly self-collected Longitudinal Set (*V*_1_, *N* = 10): 10 healthy volunteers each wore a 24-hour Holter on three days (Monday as a baseline workday, Wednesday at peak weekly workload, and Saturday as a recovery day) to test robustness across contrasting physiological contexts. (2) A Clinical Validation Set (*V*_2_, *N* = 26) from the `cded` database, comprising records with complete simultaneous ABP and ECG during standardized Valsalva and Head-Up Tilt tests; the 14 `cded` subjects in Group E were drawn exclusively from records lacking ABP signals and are non-overlapping with *V*_2_.

The *V*_1_ collection was approved by the IEC of Zhongda Hospital (No. 2019ZDSYLL073-P01). Written informed consent was obtained from all participants.

### B. Signal Preprocessing and Artifact Rejection

For datasets containing raw Electrocardiograms (ECG), signals were sampled at native resolutions (see Table I) and resampled to 250 Hz for consistency. We applied a 4th-order Butterworth bandpass filter (5–35 Hz) to remove baseline wander and high-frequency noise. R-peaks were detected using the Pan-Tompkins algorithm [18]. The resulting RR-interval time series underwent a two-stage beat-to-beat artifact rejection:

#### 1) Range Filter

Intervals outside the physiological range of [300, 2000] ms were removed to eliminate obvious technical errors.

#### 2) Quotient Filter

To rigorously exclude ectopic beats, a sliding window of 5 beats was used. An interval *RR*_*i*_ was rejected if it deviated by *>* 20% from the local median (*RR*_med_):

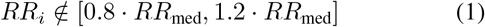

The *±*20% threshold follows the established quotient filter criterion [17], which reliably separates ectopic beats from sinus arrhythmia in long-term recordings.

This filtering process resulted in the removal of an average of 2.7% of beats per recording (representing ectopic burdens), yielding clean sinus rhythm series for feature space construction.

#### 3) Subject-Level Exclusion Criteria

Quality control was subsequently enforced at the recording level. Recordings exhibiting *>* 5% signal loss or a cumulative artifact burden exceeding 15% (based on the filters above) were designated for exclusion. In this study, no subjects were excluded, maintaining the final cohort at *N* = 209.

### C. Phase-Rectified Signal Averaging (PRSA)

The original DC proposed by Bauer et al. [4] is computed over an entire 24-hour recording and yields a single prognostic scalar. To obtain a time-resolved estimate suitable for trajectory construction, we compute DC within a 5-minute sliding window (step = 1 min), hereafter referred to as DC_local_. Within each window, PRSA aligns the RR series at all anchor points where heart rate decelerates (*RR*_*t*_ *> RR*_*t™*1_) and averages the surrounding beats. The local DC is then:

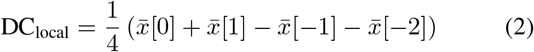

where 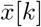 denotes the mean of all RR intervals at offset *k* relative to the anchor points (*k ∈ {−* 2, *−* 1, 0, 1}, with *k* = 0 the anchor beat). The sliding window preserves the Bauer formula’s physiological meaning while providing the temporal resolution needed for trajectory construction.

### D. HR-DC Feature Space and State Definition

HR and DC represent functionally independent dimensions of autonomic regulation: HR reflects metabolic demand and net sympatho-vagal balance, while DC quantifies vagal modulation capacity via PRSA [4]. This pairing differs from composite metrics such as SDNN or LF/HF, which aggregate sympathetic and parasympathetic contributions into a single value and cannot isolate which branch is impaired. HR and DC instead correspond to distinct physiological axes—load and vagal reserve—so their combination spans the two degrees of freedom most relevant to autonomic assessment. The two variables are only weakly correlated under most physiological conditions, and their joint value reveals whether elevated HR co-occurs with preserved or depleted vagal reserve, a distinction unavailable from either variable alone. Furthermore, a static joint distribution describes only where the system resides at a given moment; the trajectory through this space additionally encodes the direction and rate of movement between operating points, which is the more direct expression of regulatory capacity. The state vector is *P*_*t*_ = [*HR*_*t*_, *DC*_*t*_]^*⊤*^.

#### 1) Motivation for Noise Isolation

Standard DC analysis is susceptible to ectopic beats (e.g., PVCs), which inflate DC values and mimic super-vagal tone. A Gaussian Mixture Model (GMM, *k* = 3) is applied to identify and isolate the high-DC Noise Area. To distinguish artifacts from genuine vagal outbursts in healthy young subjects, a physiological continuity check is applied: valid high-DC states must persist for *≥* 3 consecutive beats, while isolated points lacking trajectory continuity are discarded.

#### 2) Individualized State Partitioning

The remaining valid space is partitioned into four physiological states. Rather than applying fixed population-wide thresholds, individualized boundaries for Deceleration Capacity (*T*_dc_) and Heart Rate (*T*_hr_) are determined from the intersection points of the GMM components. This defines Rest and Active states relative to each individual’s own physiological baseline:

- State 1 (*S*_1_): *DC ≥ T*_dc_ *∩ HR < T*_hr_.
- State 2 (*S*_2_): *DC ≥ T*_dc_ *∩ HR ≥ T*_hr_.
- State 3 (*S*_3_): *DC < T*_dc_ *∩ HR < T*_hr_.
- State 4 (*S*_4_): *DC < T*_dc_ *∩ HR ≥ T*_hr_.

The complete framework is summarized in Fig. 1.

**Fig. 1.**
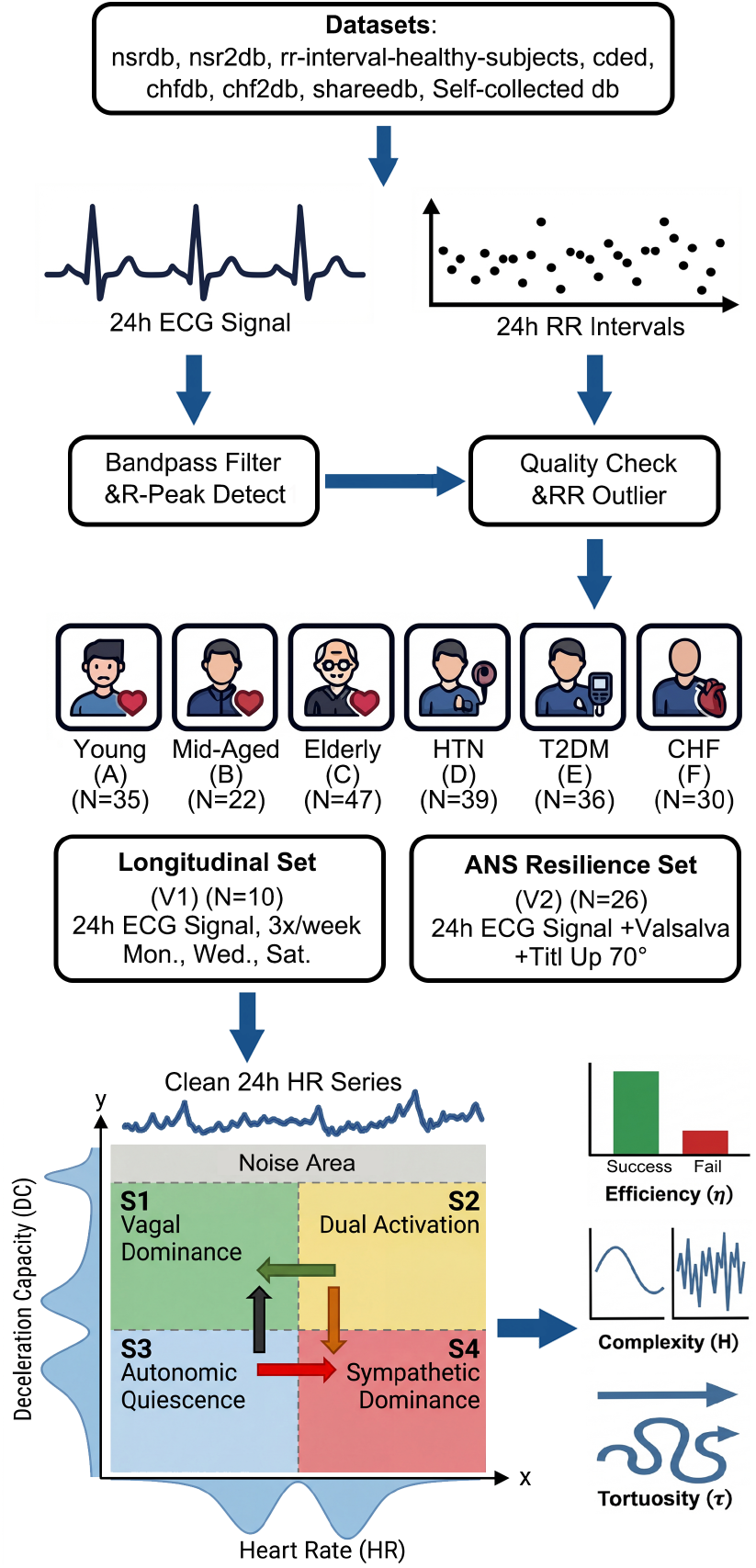
Overview of the Study Framework. (Top) Data acquisition from heterogeneous sources, preprocessing pipeline, and stratification of the main study cohort (*N* = 209, Groups A–F) and two validation subsets (*V*_1_, *V*_2_). (Bottom Left) HR-DC feature space construction. The GMM partitions the space into four physiological states (*S*_1_–*S*_4_) while explicitly isolating the high-DC Noise Area. The colored arrows represent extracted functional dynamics: *Relaxation Stream* (Green) and *Excitation Stream* (Red). (Bottom Right) Derivation of the three dynamic biomarkers: Transition Efficiency (*η*), Directional Complexity (*H*), and Tortuosity (*τ*).

### E. Bidirectional Trajectory Extraction

Trajectories were extracted for transitions *from* intermediate states (*S*_2_, *S*_3_) *towards* two target states. *S*_1_ (high DC, low HR) is the Relaxation target, representing restored vagal tone at reduced metabolic load. *S*_4_ (low DC, high HR) is the Excitation target, representing sustained sympathetic mobilization in which vagal modulation is physiologically suppressed to support elevated heart rate demand, as occurs during exertion or acute stress. Path A (*S*_2_ *→ S*_1_) and Path B (*S*_3_ *→ S*_1_) constitute the Relaxation stream; Path C (*S*_2_ *→ S*_4_) and Path D (*S*_3_ *→ S*_4_) constitute the Excitation stream. A valid trajectory under the Onset-Track-Stabilize protocol requires a stable dwell (*T*_dwell_ = 10 min) in the source state and a stabilization window (*W*_stab_ = 5 min) in the target. Trajectories that fail to stabilize within *T*_max_ = 40 min are discarded; this window aligns with the semi-period of the ultradian Basic Rest-Activity Cycle and filters out prolonged, undirected fluctuations.

### F. Metrics Definition

Three metrics are computed for two streams. The Relaxation stream pools all valid trajectories from Paths A and B; the Excitation stream pools Paths C and D. Aggregation is at the trajectory level, so stream-level values reflect the combined set rather than a simple average of two path-level values.

#### 1) Transition Efficiency (η)

The probability of successfully reaching the target state:

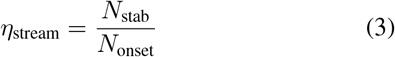

where *N*_stab_ is the count of successfully stabilized transitions and *N*_onset_ is the total number of detected onsets.

#### 2) Directional Complexity (H)

Shannon entropy of the pooled trajectory angle distribution, quantifying control smoothness:

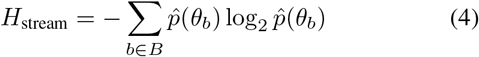

where *B* is the set of 18 directional bins (20° each), and 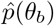 is estimated by pooling all inter-sample step angles from the complete set of valid trajectories *T* in the stream. Pooling yields a single well-defined distribution regardless of individual trajectory length.

#### 3) Directional Tortuosity (τ)

Because HR (bpm) and DC (ms) carry different physical units, the feature space axes are z-score normalized per subject before any distance computation, using the mean and standard deviation of all valid *P*_*t*_ points in that recording. This ensures that *τ* is dimensionless and invariant to the choice of physical units. The arc-chord ratio is then:

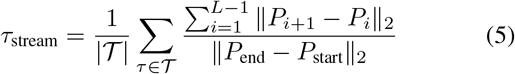

where *∥ · ∥*_2_ denotes the Euclidean distance in the normalized space, *P*_start_ is the onset point, and *P*_end_ is the stabilization point.

### G. Statistical Analysis and Machine Learning

Data are expressed as mean *±* standard deviation. Statistical significance was defined as *p <* 0.05.

- Power Analysis: A post-hoc power analysis was conducted using G*Power 3.1. For a one-way ANOVA with 6 groups and a total sample size of *N* = 209, assuming a medium-to-large effect size (Cohen’s *f* = 0.35) and *α* = 0.05, the calculated statistical power (1 *™ β*) exceeded 0.98. This confirms the dataset is sufficient to detect significant group differences.
- Hypothesis Testing: Group differences were assessed using One-way ANOVA followed by Bonferroni post-hoc corrections for two planned comparisons: (1) Ageing effects (Groups A–C) and (2) Pathological deviations relative to healthy elderly controls (Groups D–F vs. C).
- Predictive Modeling Framework: To evaluate the diagnostic and translational utility of the proposed biomarkers, we implemented a comparative machine learning framework using 5-fold cross-validation. The input feature vector consisted of the six derived metrics ([*η, H, τ*]_*rel*_, [*η, H, τ*]_*exc*_). To prevent data leakage, all feature normalization (Z-score scaling) was performed strictly within each cross-validation loop. Parameters (mean and standard deviation) were derived solely from the training folds and applied to the test fold.
  - Physiological State Classification: To discriminate Healthy Elderly from Pathological subjects (HTN+T2DM+CHF), we benchmarked Support Vector Machine (SVM, Gaussian Kernel), Random Forest, and Logistic Regression. Performance was quantified using the AUC and Accuracy.
  - Clinical Metrics Regression: To predict physiological ground truths, we compared Robust Linear Regression, Ridge Regression, and Support Vector Regression (SVR). Performance was evaluated using *R*^2^ and RMSE.

## III. Results

### A. Robustness: Decoupling Capacity from State

Analysis of the Longitudinal Set (*N* = 10) across three days (Mon/Wed/Sat) is summarized in Table II. Mean HR rose significantly on the high-workload day (Wed: 78 *±* 6 bpm vs. Sat: 67 *±* 5 bpm, *p <* 0.01), and Mean DC showed a compensatory decrease (4.8 *±* 1.1 vs. 7.5 *±* 1.5 ms, *p <* 0.01). The proposed metrics (*η, H, τ*) showed no significant variation across days (*p >* 0.05, *ICC >* 0.85), indicating they reflect a stable individual characteristic rather than a transient state.

**TABLE 2.**
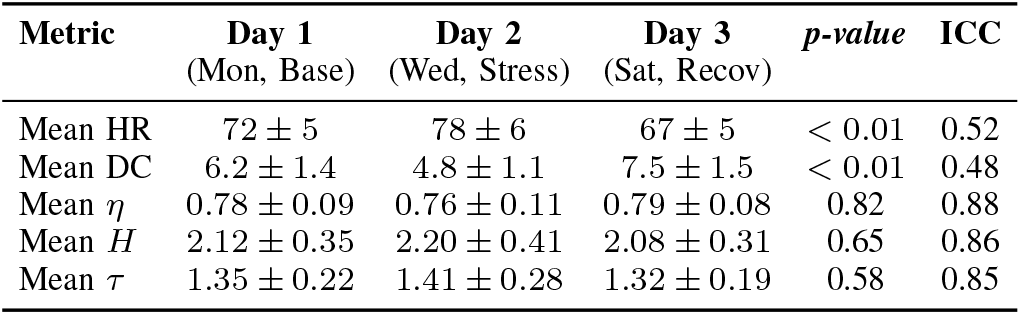
Test-Retest Reliability Across Contexts (N=10)

| Metric | Day 1<br>(Mon, Base) | Day 2<br>(Wed, Stress) | Day 3<br>(Sat, Recov) | p-value | ICC |
| --- | --- | --- | --- | --- | --- |
| Mean HR | $72 \pm 5$ | $78 \pm 6$ | $67 \pm 5$ | $< 0.01$ | 0.52 |
| Mean DC | $6.2 \pm 1.4$ | $4.8 \pm 1.1$ | $7.5 \pm 1.5$ | $< 0.01$ | 0.48 |
| Mean $\eta$ | $0.78 \pm 0.09$ | $0.76 \pm 0.11$ | $0.79 \pm 0.08$ | 0.82 | 0.88 |
| Mean $H$ | $2.12 \pm 0.35$ | $2.20 \pm 0.41$ | $2.08 \pm 0.31$ | 0.65 | 0.86 |
| Mean $\tau$ | $1.35 \pm 0.22$ | $1.41 \pm 0.28$ | $1.32 \pm 0.19$ | 0.58 | 0.85 |

### B. Feature Space Topology and Trajectory Dynamics

Fig. 2 shows aggregate feature space densities and representative trajectories for each group. Healthy subjects (A– C) exhibit a characteristic L-shaped topology with smooth, ballistic trajectories between the Rest (*S*_1_) and Active (*S*_4_) regions. In HTN (D), the recovery arm is truncated: trajectories frequently initiate from *S*_2_ but stall before reaching *S*_1_, indicating impaired vagal withdrawal. T2DM (E) produces a diffuse, globular cloud with erratic, high-frequency trajectory fluctuations rather than directional state transitions. In CHF (F), the feature space collapses to a dense cluster near the origin with extremely short, confined trajectories and near-complete loss of dynamic range.

**Fig. 2.**
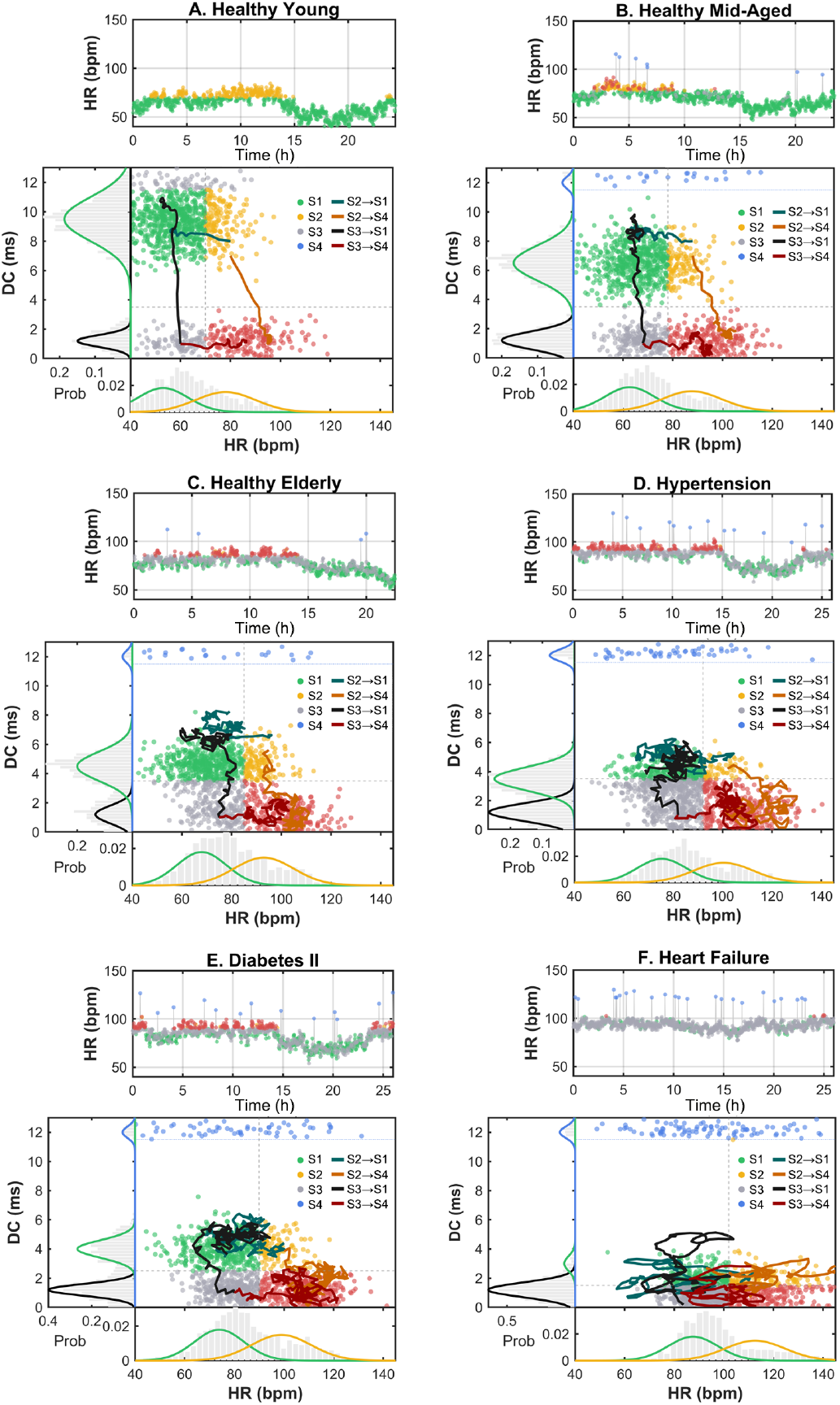
HR-DC Feature Space Topology and Trajectory Characteristics. Representative subjects from each group are shown. (A–C) Healthy aging preserves the L-shaped topology with smooth, ballistic trajectories. (D) Hypertension shows a truncated recovery arm with stalled trajectories. (E) Type 2 Diabetes exhibits a diffuse cloud with erratic, high-frequency trajectory fluctuations. (F) Congestive Heart Failure exhibits a collapsed, low-range cluster with extremely short trajectories.

### C. Quantitative Population Stratification

Table III-C2 reports the proposed metrics across all six groups.

#### 1) Age-Dependency

The framework detected significantly reduced Efficiency (*η*) and increased Tortuosity (*τ*_*rel*_) already in Group B relative to Group A (*p <* 0.05), a separation that is not reliably captured by traditional HRV metrics in the middle-aged range. Group C showed further decline (*p <* 0.01), consistent with progressive loss of homeostatic range with age.

#### 2) Pathological Deviations

Compared to age-matched Healthy Elderly (Group C), HTN exhibited significantly lower *η*_*rel*_ (*p <* 0.01) with preserved *η*_*exc*_ (*p >* 0.05), a pattern of asymmetric impairment consistent with the sympatho-vagal imbalance seen in baroreflex resetting. T2DM showed significantly elevated Complexity (*H*) and Tortuosity (*τ*) in both streams (*p <* 0.05), reflecting the tremulous, multidirectional trajectory fluctuations observed qualitatively in Fig. 2E. CHF produced the lowest efficiency scores across all metrics (*η ≤* 0.32, *p <* 0.001), indicating near-total suppression of directed autonomic maneuvers.

### D. Physiological and Clinical Validation

To validate physiological relevance, we analyzed the ANS Clinical Set (*V*_2_, *N* = 26) containing active reflex data.

#### 1) Ground Truth Calculation (Fig. 3A, B)

We derived gold-standard indices from raw signals:

**Fig. 3.**
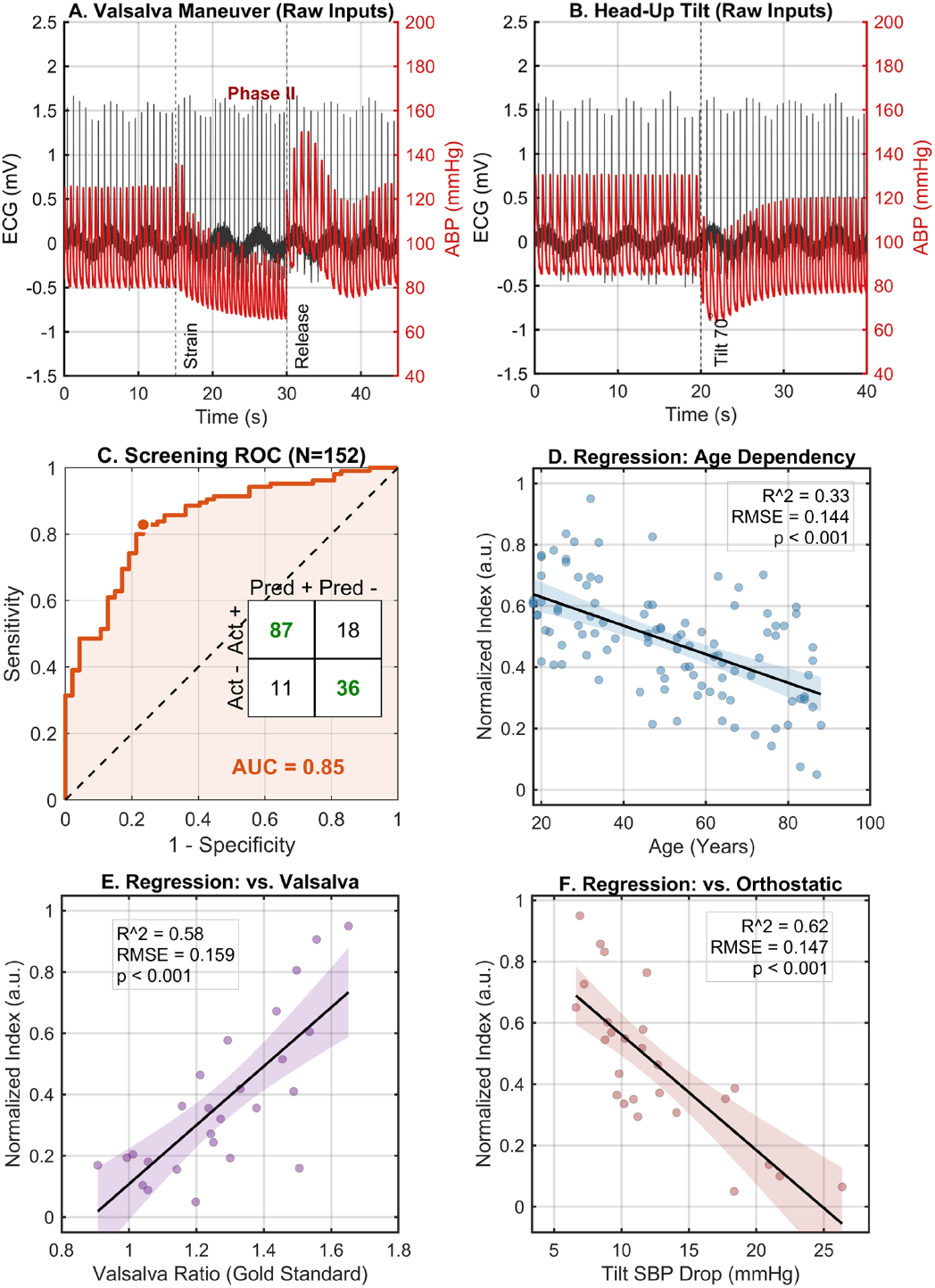
Physiological and Clinical Validation. (A) Raw ECG and ABP during active Valsalva Maneuver. (B) Raw signals during Tilt Test showing SBP drop calculation. (C) ROC Curve for SVM classifier using combined metrics (Healthy Elderly vs Pathology, AUC=0.85). (D) Age-dependency regression. (E) Multivariate Regression: Passive Metrics (*η*_*rel*_, *H*_*rel*_, *τ*_*rel*_) vs Active Valsalva Ratio (*R*^2^ = 0.58). (F) Multivariate Regression: Passive Metrics (*η*_*exc*_, *H*_*exc*_, *τ*_*exc*_) vs Tilt SBP Drop (*R*^2^ = 0.62).

- Valsalva Ratio (VR): Calculated from the standard Valsalva Maneuver (40mmHg intrathoracic pressure for 15s).

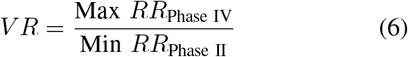

Phase II represents sympathetic compensation (tachycardia) during strain, while Phase IV represents the vagal rebound (bradycardia) upon release.

- Tilt SBP Drop: The maximum systolic blood pressure drop during the 10-min Head-Up Tilt (70°).

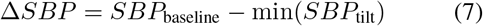

This quantifies sympathetic vasomotor failure; a larger drop indicates poorer vasomotor tone, and subjects with higher *η*_*exc*_ are expected to show smaller Δ*SBP* .

| Grp | Relaxation Stream ($\rightarrow S_1$) | | | Excitation Stream ($\rightarrow S_4$) | | |
| --- | --- | --- | --- | --- | --- | --- |
| | $\eta_{\text{rel}}$ | $H_{\text{rel}}$ | $\tau_{\text{rel}}$ | $\eta_{\text{exc}}$ | $H_{\text{exc}}$ | $\tau_{\text{exc}}$ |
| A | 0.87 (0.07) | 1.82 (0.22) | 1.15 (0.12) | 0.84 (0.08) | 1.88 (0.24) | 1.18 (0.15) |
| B | 0.81 (0.09)* | 1.92 (0.28) | 1.24 (0.18)* | 0.79 (0.10)* | 1.96 (0.30) | 1.25 (0.20) |
| C | 0.76 (0.14)** | 2.05 (0.35)* | 1.38 (0.25)** | 0.75 (0.15)* | 2.10 (0.38)* | 1.42 (0.28)** |
| HTN | 0.60 (0.15)†† | 2.22 (0.40) | 1.65 (0.35)† | 0.73 (0.16) | 2.18 (0.42) | 1.55 (0.32) |
| T2DM | 0.65 (0.22)† | 2.45 (0.60)†† | 2.10 (0.55)†† | 0.62 (0.24)† | 2.52 (0.65)† | 2.20 (0.60)†† |
| CHF | 0.30 (0.18)††† | 2.15 (0.65) | 1.95 (0.85) | 0.32 (0.20)††† | 2.20 (0.70) | 2.05 (0.90) |

Values: Mean (SD). Age rows (A–C): \* $p < 0.05$ , \*\* $p < 0.01$ vs Grp A. Disease rows (D–F): † $p < 0.05$ , †† $p < 0.01$ , ††† $p < 0.001$ vs Grp C.

#### 2) Physiological State Classification (Fig. 3C)

Using the combined feature vector [*η, H, τ*], the SVM (Gaussian Kernel) outperformed Random Forest and Logistic Regression, achieving an AUC of 0.85 (Accuracy: 80.9%).

#### 3) Clinical Metrics Regression (Fig. 3E, F)

Robust Linear Regression using [*η, H, τ*] as predictors showed that Relaxation metrics predicted the Valsalva Ratio (*R*^2^ = 0.58, *p <* 0.001) and Excitation metrics predicted the Tilt SBP Drop (*R*^2^ = 0.62, *p <* 0.001), providing evidence for stream-specific correspondence between passive trajectory dynamics and active vagal and sympathetic reflex responses, respectively.

## IV. Discussion

### A. Beyond Static Variability

Conventional metrics (Mean HR, Mean DC) fluctuated substantially across days (*p <* 0.01), while *η, H*, and *τ* remained stable (*ICC >* 0.85), indicating that trajectory-based metrics reflect a stable individual property rather than a transient state. The framework yields six scalar biomarkers from two subject-specific thresholds (*T*_dc_, *T*_hr_), a dimensionality consistent with symmetry constraints in a two-variable dynamical system [16].

### B. Pathological Phenotyping

The three disease groups produced qualitatively different impairment patterns. HTN selectively impaired the Relaxation stream while sparing Excitation, consistent with baroreflex resetting that preserves sympathetic reactivity while shifting the cardiovascular operating point [19]. T2DM elevated both *H* and *τ* globally, reflecting the stochastic, multidirectional trajectory fluctuations associated with cardiac autonomic neuropathy and neural temporal dispersion [20]. CHF produced global suppression of *η* across both streams, consistent with end-stage depletion of neurotransmitter reserves and receptor downregulation [21]. That each condition produces a distinct metric fingerprint suggests the framework may complement diagnosis beyond a binary healthy/pathological classification.

### C. Correspondence with Active Reflex Tests and Future Directions

The correlation between passive trajectory metrics and active reflex outcomes (*R*^2^ = 0.58 for Valsalva, *R*^2^ = 0.62 for Tilt) suggests that spontaneous recovery events during normal 24-hour activity share underlying neural pathways with responses to forced maneuvers, and could enable reflex assessment in patients unable to cooperate with active testing. The GMM-based Noise Area also filters sensor artifacts spatially in the HR-DC plane, suggesting the algorithm may tolerate noisier RR estimates from wrist-worn photoplethysmography; validation on PPG data is a natural next step.

## V. Conclusion

We have described HR-DC State-Transition Dynamics, a method for extracting directed state-transition metrics from 24-hour RR interval recordings. The proposed biomarkers (*η, H, τ*) showed high test-retest reliability across contrasting activity contexts (*ICC >* 0.85), separated three pathological groups from age-matched healthy controls, and correlated with active Valsalva and tilt test outcomes (*R*^2^ = 0.58 and 0.62), suggesting the framework can characterize autonomic regulatory function from routine monitoring without active patient cooperation.

## Data Availability

All data produced in the present study are available upon reasonable request to the authors.

## VI. Acknowledgment

We gratefully acknowledge the PhysioNet team for providing open-access physiological data. We also thank the investigators and staff involved in the original data collection, and most sincerely, the participants who contributed to these studies.

## Notes

### Competing Interest Statement

The authors have declared no competing interest.

### Author Declarations

The Independent Ethics Committee for Clinical Research of Zhongda Hospital, affiliated with Southeast University, gave ethical approval for this work (approval No. 2019ZDSYLL073-P01). Participants in the internally collected cohorts provided written informed consent. The Institutional Review Board of Beth Israel Deaconess Medical Center approved the original collection of the publicly available cded dataset (approval No. 2008P000286).

### Funding

- National Natural Science Foundation of China: 62477007

